## Supplementary Material for "Spatiotemporal dynamics and epidemiological impact of SARS-CoV-2 XBB lineages dissemination in Brazil in 2023"

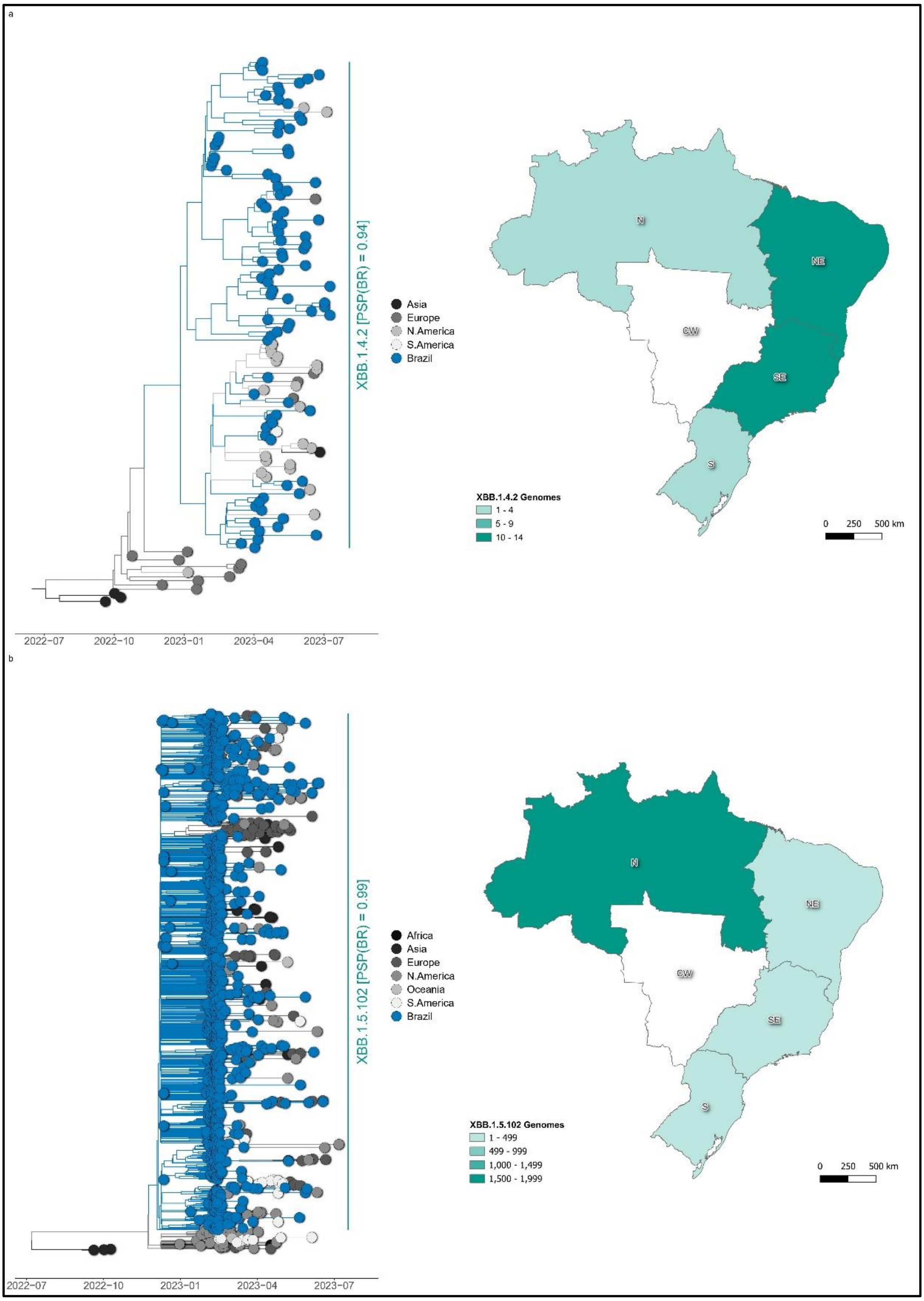


***Supplementary Figure 1: Time-scaled phylogeographic trees of SARS-CoV-2 XBB* lineages. a.*** *XBB.1.4.2 (n = 131),* ***b.*** *XBB.1.5.102 (n = 1,274). The trees are colored in accordance with the reconstructed location of their internal nodes and following the color code on the right side of each panel. Key lineages in each tree are named by labels on the right side of each panel, which also contains their posterior state probability of emergence in Brazil. Each of these trees is accompanied by maps illustrating their prevalence across Brazil's five geographical regions: N (North), NE (Northeast), CW (Central-West), SE (Southeast), and S (South). Maps were generated with QGIS v.3.10.2 software (http://qgis.org) using public access data downloaded from the GADM v.3.6 database (https://gadm.org) and shapefiles obtained from the Brazilian Institute of Geography and Statistics (https://portaldemapas.ibge.gov.br/portal.php#homepage).*

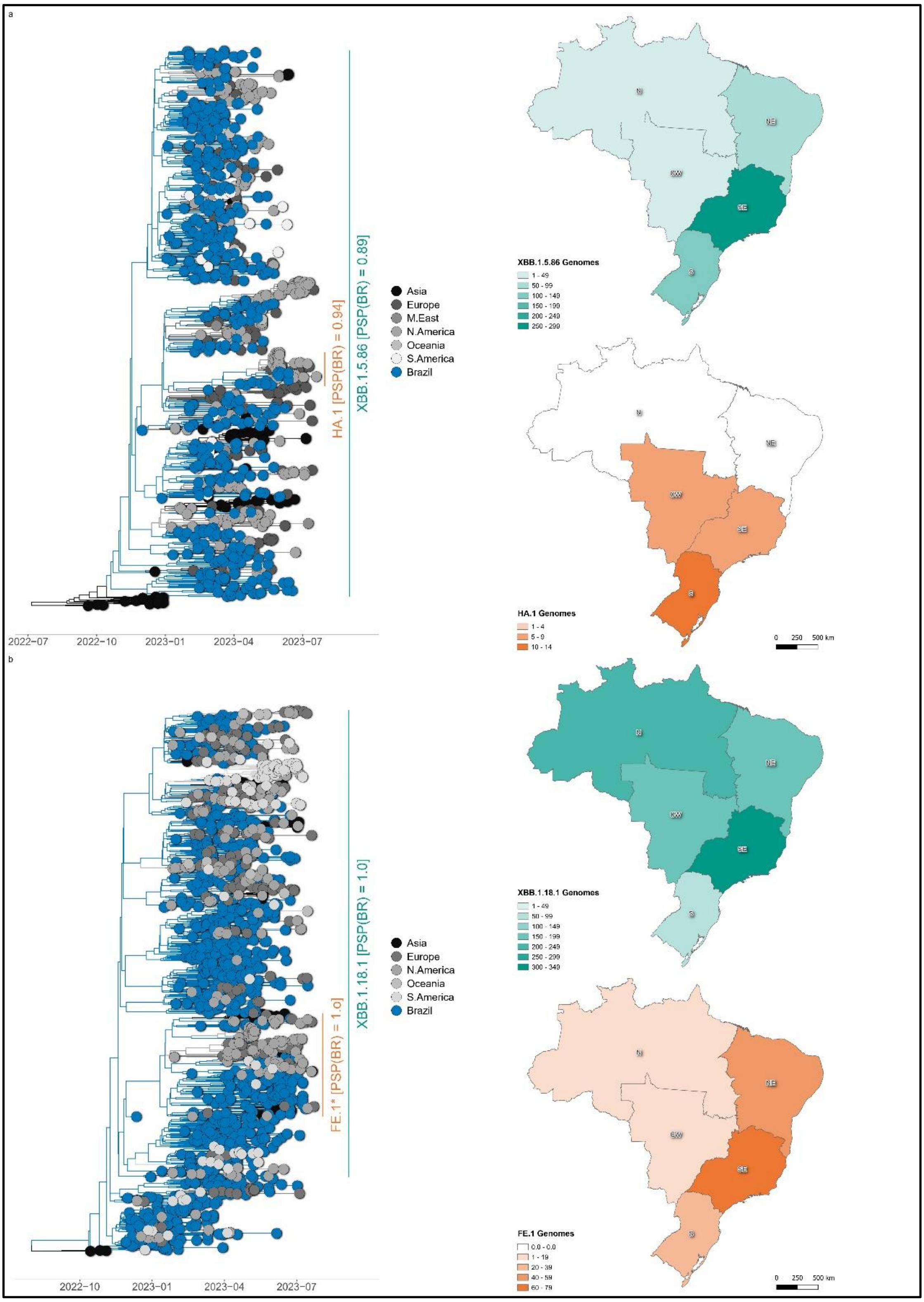


***Supplementary Figure 2: Time-scaled phylogeographic trees of SARS-CoV-2 XBB* lineages. a.*** *XBB.1.5.86/HA.1 (n = 991),* ***b.*** *XBB.1.18.1/FE.1 (n = 1,544). The trees are colored in accordance with the reconstructed location of their internal nodes and following the color code on the right side of each panel. Key lineages in each tree are named by labels on the right side of each panel, which also contains their posterior state probability of emergence in Brazil. Each of these trees is accompanied by maps illustrating their prevalence across Brazil's five geographical regions: N (North), NE (Northeast), CW (Central-West), SE (Southeast), and S (South). Maps were generated with QGIS v.3.10.2 software (http://qgis.org) using public access data downloaded from the GADM v.3.6 database (https://gadm.org) and shapefiles obtained from the Brazilian Institute of Geography and Statistics (https://portaldemapas.ibge.gov.br/portal.php#homepage).*

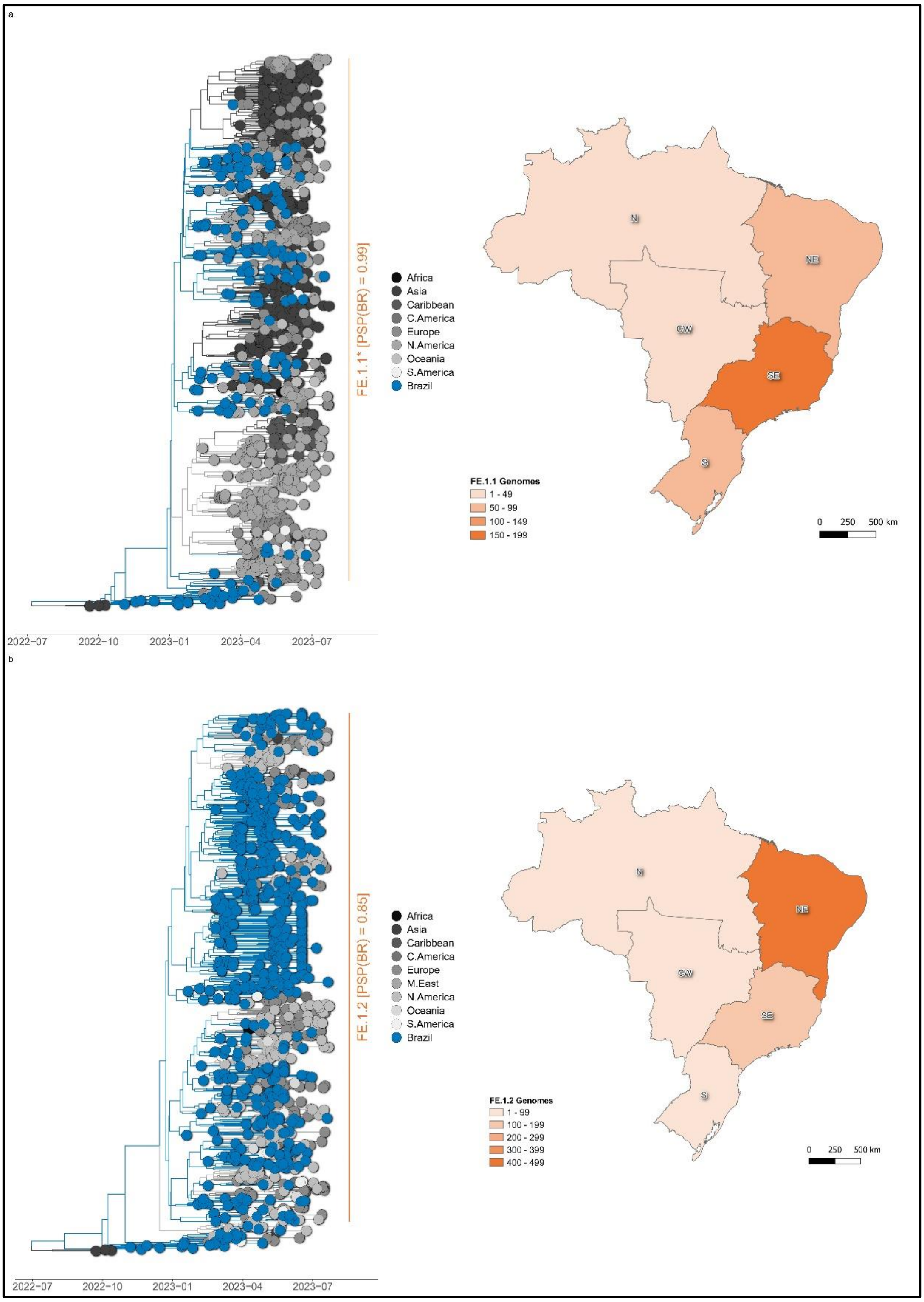


***Supplementary Figure 3: Time-scaled phylogeographic trees of SARS-CoV-2 XBB* lineages*. a.** *FE.1.1 (n = 1,251),* ***b.*** *FE.1.2 (n = 1,047). The trees are colored in accordance with the reconstructed location of their internal nodes and following the color code on the right side of each panel. Key lineages in each tree are named by labels on the right side of each panel, which also contains their posterior state probability of emergence in Brazil. Each of these trees is accompanied by maps illustrating their prevalence across Brazil's five geographical regions: N (North), NE (Northeast), CW (Central-West), SE (Southeast), and S (South). Maps were generated with QGIS v.3.10.2 software (http://qgis.org) using public access data downloaded from the GADM v.3.6 database (https://gadm.org) and shapefiles obtained from the Brazilian Institute of Geography and Statistics (https://portaldemapas.ibge.gov.br/portal.php#homepage).*

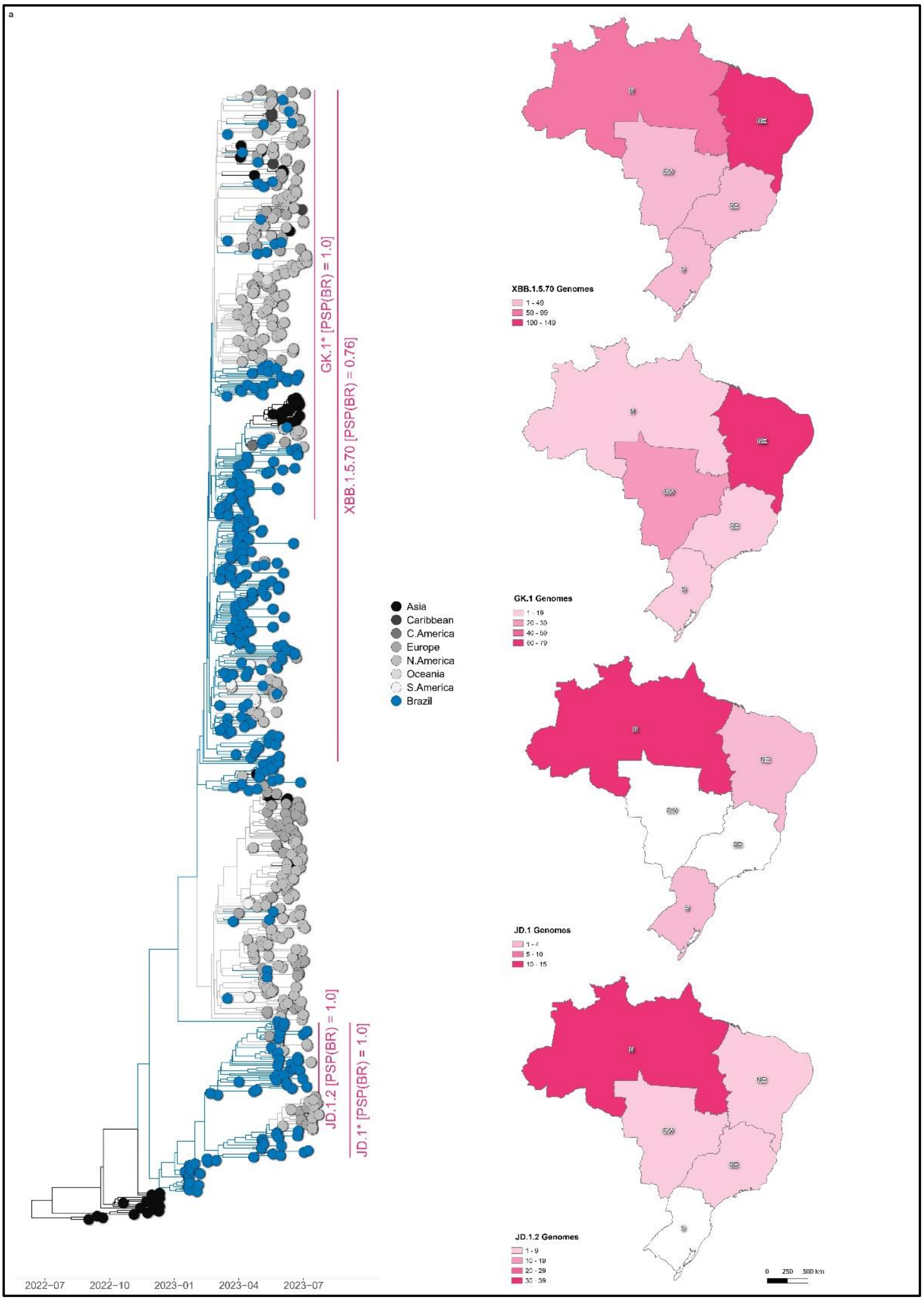


***Supplementary Figure 4: Time-scaled phylogeographic trees of SARS-CoV-2 XBB* lineages*. a.** *XBB.1.5.70/GK.1*/JD.1*/JD.1.2 (n = 691). The tree is colored in accordance with the reconstructed location of its internal nodes and following the color code on the right side of the panel. Key lineages in each tree are named by labels on the right side of each panel, which also contains their posterior state probability of emergence in Brazil. Each of these trees is accompanied by maps illustrating their prevalence across Brazil's five geographical regions: N (North), NE (Northeast), CW (Central-West), SE (Southeast), and S (South). Maps were generated with QGIS v.3.10.2 software (http://qgis.org) using public access data downloaded from the GADM v.3.6 database (https://gadm.org) and shapefiles obtained from the Brazilian Institute of Geography and Statistics (https://portaldemapas.ibge.gov.br/portal.php#homepage).*

**Supplementary Table 1:** Major Brazilian SARS-CoV-2 Omicron XBB* lineages.

| XBB Group | Pango Lineage | N | Sampling Range | $T_{MRCA}$ (95% HPD) | aLRT | Root (*PSP*) |
|---|---|---|---|---|---|---|
| XBB*+F486P | XBB.1.5.102 | 1,226 | 2022-12-09/ 2022-07-08 | 2022-12-03 (2022-11-22/2022-12-08) | 0.89 | BR (0.99) |
| | XBB.1.18.1 | 936 | 2022-11-10/ 2023-07-16 | 2022-10-17 (2022-10-02/2022-10-28) | 0.85 | BR (1.0) |
| | XBB.1.5.86 | 877 | 2022-12-01/ 2023-07-15 | 2022-11-05 (2022-10-13/2022-11-22) | 0.83 | BR (0.89) |
| | XBB.1.4.2 | 118 | 2023-02-05/ 2023-07-10 | 2022-12-29 (2022-11-30/2023-01-21) | 0.94 | BR (0.94) |
| XBB*+F486P+F456L | FE.1 (XBB.1.18.1.1) | 152 | 2023-01-26/ 2023-07-17 | 2022-12-12 (2022-11-24/2022-12-25) | 1.0 | BR (1.0) |
| | FE.1.1 (XBB.1.18.1.1.1) | 1,193 | 2023-02-02/ 2023-07-24 | 2022-12-18 (2022-11-26/2023-01-04) | 0.89 | BR (0.99) |
| | FE.1.2 (XBB.1.18.1.1.2) | 990 | 2022-01-16/ 2023-07-21 | 2022-12-21 (2022-12-04/2023-01-04) | 0.85 | BR (1.0) |
| | HA.1 (XBB.1.5.86.1) | 60 | 2023-04-10/ 2023-07-21 | 2023-01-24 (2023-01-07/2023-02-13) | 0.85 | BR (0.94) |
| XBB*+F486P+F456L+L455F | XBB.1.5.70 | 214 | 2023-03-24/ 2023-07-21 | 2023-02-14 (2023-01-30/2023-02-27) | 1.0 | BR (0.76) |
| | GK.1 (XBB.1.5.70.1) | 148 | 2023-03-24/ 2023-07-22 | 2023-03-09 (2023-03-03/2023-03-15) | 0.78 | BR (1.0) |
| | GK.1.1 (XBB.1.5.70.1.1) | 36 | 2023-05-09/ 2023-07-20 | 2023-04-06 (2023-03-18/2023-04-22) | 0.92 | BR (1.0) |
| | GK.1.2 (XBB.1.5.70.1.2) | 12 | 2023-05-21/ 2023-07-18 | 2023-04-16 (2023-03-19/2023-05-03) | 0.93 | NA (0.90) |
| | GK.1.3 (XBB.1.5.70.1.3) | 67 | 2023-04-18/ 2023-07-25 | 2023-03-19 (2023-03-12/2023-03-25) | 0.96 | NA (1.0) |
| | GK.2 (XBB.1.5.70.2) | 68 | 2023-05-05/ 2023-07-18 | 2023-04-13 (2023-03-26/2023-04-28) | 0.91 | NA (0.70) |
| | GK.3 (XBB.1.5.70.3) | 21 | 2023-05-18/ 2023-07-02 | 2023-04-14 (2023-03-25/2023-04-30) | 0.87 | NA (1.0) |
| | JD.1 (XBB.1.5.102.1) | 19 | 2023-05-11/ 2023-07-31 | 2023-03-02 (2023-02-20/2023-03-09) | 0.97 | BR (1.0) |
| | JD.1.1 (XBB.1.5.102.1.1) | 19 | 2023-06-21/ 2023-07-31 | 2023-06-14 (2023-06-06/2023-06-20) | 0.83 | NA (0.93) |
| | JD.1.2 (XBB.1.5.102.1.2) | 45 | 2023-03-11/ 2023-07-31 | 2023-03-09 (2023-03-04/2023-03-10) | 0.90 | BR (1.0) |

*The table details the space-time dynamics of autochthonous XBB* Brazilian lineages and their descendants. For better comprehension of their ancestry, selected lineages have been partially aliased. All lineages are accompanied by (i) the number of sequences in the phylogeographic reconstruction; (ii) sampling range; (iii)* $T_{MRCA}$ *and corresponding 95% HPD; (iv) statistical support (aLRT) in the original maximum likelihood topology; (v) most probable location of their MRCA, and its statistical support (PSP). NA: North America, BR: Brazil.*

### COVID-19 FIOCRUZ GENOMIC SURVEILLANCE NETWORK MEMBERS

**FIOCRUZ PERNAMBUCO**

***Instituto Aggeu Magalhães***

Gabriel da Luz Wallau
Marcelo Henrique Santos Paiva
Roberto Dias Lins Neto
Cassia Docena
Matheus Filgueira Bezerra
Alexandre Freitas da Silva
Danilo Fernandes Coêlho
Duschinka Ribeiro Duarte Guedes
Emerson Gonçalves Moreira
Filipe Zimmer Dezordi
Gustavo Barbosa de Lima
Isabelle Freire Viana
Lais Ceschini Machado
Lilian Caroliny Amorim Silva
Marcus André Molinero
Matheus Vitor Ferreira Ferraz
Raul Emidio de Lima
Tayná Evily de Lima
Tulio de Lima Campos
Wenny Camilla dos Santos Adan

**FIOCRUZ BAHIA**

***Instituto Gonçalo Moniz***

Antônio Ricardo Khouri Cunha
Bruno de Bezerril Andrade
Bruno Solano de Freitas Souza
Camila Indiani de Oliveira
Clarissa Araújo Gurgel Rocha
Isadora Cristina de Siqueira
Leonardo Paiva Farias
Vanessa Leiko Oikawa Cardoso

**FIOCRUZ RIO DE JANEIRO**

***Instituto Oswaldo Cruz***

***Laboratório de Vírus Respiratórios, Exantemáticos, Enterovírus e Emergências Virais***

Marilda Teixeira de Siqueira
Paola Cristina Resende Silva
Fernando do Couto Motta
Alice Sampaio Barreto da Rocha
Bruna Mendonça da Silva
Edson Elias da Silva
Elisa Cavalcante Pereira
Gabriela Calegario
Jéssica Graça de Macedo Carvalho
Larissa Macedo Pinto
Leonardo Saboia Vahia Matilde
Lucas Freitas
Luciana Appolinario
Renata Serrano Lopes
Victor Guimarães Ribeiro

***Laboratório de Arbovírus e Vírus Hemorrágicos***

Ana Maria Bispo de Filippis
Felipe Gomes Naveca
Gonzalo Bello
Marcos César Lima de Mendonça
Ighor Leonardo Arantes Gomes

***Laboratório de Genômica Funcional e Bioinformática***

Wim M. S. Degrave

**Plataforma Genômica**

Aline dos Santos Moreira
Alexandre Cunha dos Santos
Audrien Alves Andrade de Souza
Beatriz de Lima Alessio Müller
Camila Castanon Freire Barraca
Marília Alves Figueira de Melo

**Plataforma de Bioinformática**

Thiago Estevam Parente
Rafael Ferreira Soares
Daniel Andrade Moreira

**FIOCRUZ AMAZÔNIA**

***Instituto Leônidas e Maria Deane***

Felipe Gomes Naveca
André de Lima Guerra Corado
Fernanda Oliveira do Nascimento
George Allan Villarouco da Silva
Karina Pinheiro Pessoa
Luciana Mara Fé Gonçalves
Maria Julia Pessoa Brandão
Matilde Contreras Mejía
Valdinete Alves do Nascimento
Victor Costa de Souza

***Instituto Nacional de Controle de Qualidade em Saúde***

Maysa Mandetta Clementino
Andressa Silva Gonçalves de Brito
Beatriz Oliveira de Farias
Ivano Victorio de Filippis Capasso
Kayo Cesar Bianco Fernandes
Mariana Magaldi de Souza Lima

**FIOCRUZ MINAS GERAIS**

***Instituto René Rachou***

Pedro Augusto Alves
Gabriel da Rocha Fernandes
Anna Christina de Matos Salim
Carlos Eduardo Calzavara Silva
Caroline Penido Rocha
Flávio Marcos Gomes de Araújo
Rubens Lima do Monte Neto
Thaís Bárbara de Souza Silva

**FIOCRUZ RONDÔNIA**

Deusilene Souza Vieira Dall'Acqua
Jackson Alves da Silva Queiroz
Ana Maisa Passos da Silva
Jessiane Rodrigues Ribeiro

**FIOCRUZ CEARÁ**

Fabio Miyajima
Fernando Braga Stehling Dias
Alice Di Sabatino Guilmarães
Antonio Lucas Delerino
Carlos Leonardo de Aragão Araújo
Cecília Leite Costa
Cleber Furtado Aksenen
Eduardo Ruback dos Santos
Igor Oliveira Duarte
Jamille Maria Mendes Bezerra
Joaquim Sousa Junior
Marcela Helena Gambim Fonseca
Nicole Silva França
Pedro Miguel Carneiro Jeronimo
Suzana Porto Almeida
Thais de Oliveira Costa
Thaís Ferreira de Oliveira
Ticiane Cavalcante de Souza
Veridiana Pessoa Miyajim

**VICE-PRESIDÊNCIA DE ENSINO, INFORMAÇÃO E COMUNICAÇÃO**

Marcelo Ferreira da Costa Gomes
Antonio Fonseca Pacheco
Claudia Torres Codeço
Daniel Antunes Maciel Villela
Leonardo Soares Bastos
Raquel Martins Lana

**FIOCRUZ PIAUÍ**

Vladimir Costa Silva
Adelino Soares Lima Neto
Carlos Henrique Nery Costa
Hérida Jeyne de Oliveira Amaral
Jacenir Reis dos Santos Mallet
Walterlene de Carvalho Gonçalves

**FIOCRUZ PARANÁ**

**Instituto Carlos Chagas**

Helisson Faoro
Tiago Gräf
Andrea Rodrigues Avila
Andréia Akemi Suzukawa
Bruno Dallagiovanna
Eduardo Balsanelli
Emanuel Maltempi de Souza
Fabio de Oliveira Pedrosa
Fabio Passetti
Fabricio Klerynton Marchini
Hellen Geremias dos Santos
Luis Gustavo Morello
Mauro Medeiros de Oliveira
Michelle Orane Schemberger
Valter Antonio de Baura
Wagner Nagib

**FIOCRUZ MATO GROSSO DO SUL**

Alexsandra Mendonça Favacho
Zoraida Fernandez Grillo
Daniel Máximo Corrêa Alcântara
Thiago Fernandes de Oliveira

**VICE-PRESIDÊNCIA E DE PESQUISA E COLEÇÕES BIOLÓGICAS**

Rodrigo Correa de Oliveira
Bruno de Sousa Moraes
Cristiane Elisa Boar
